# Addressing inequalities in vaccination coverage among children aged 12 to 23 months in ten Sub-Saharan African countries: Insights from DHS and MIS Data (2017-2022)

**DOI:** 10.1101/2024.12.13.24318976

**Authors:** Sancho Pedro Xavier, Manuel Mahoche, Patrícia Helen Rondó, Ageo Mário da Silva, Renzo Flores-Ortiz, Audêncio Victor

**Author notes:** **Correspondence:** Sancho Pedro Xavier, Federal University of Mato Grosso - Institute of Collective Health, Cuiaba-MT, 78060-900, Brazil. Equal contribution.

## Abstract

**Introduction:** Vaccination is one of the most effective public health interventions for preventing and controlling infectious diseases, particularly in low- and middle-income countries. This study analyzed disparities in vaccination coverage among children aged 12 to 23 months in ten Sub-Saharan African (SSA) countries.

**Methods:** A cross-sectional study using data from Demographic and Health Surveys (DHS) and Multiple Indicator Cluster Surveys (MICS) collected between 2017 and 2022 in ten Sub-Saharan African countries. The primary outcome was full vaccination coverage. Logistic regression models were applied to identify factors associated with the outcome.

**Results:** Full vaccination coverage rates varied significantly across countries, with Gambia presenting the highest rate (86.4%) and Guinea the lowest (21.2%). Factors associated with full vaccination coverage included place of residence, maternal education and age, economic status, place of delivery, possession of a health card, and attendance at prenatal care services.

**Conclusion:** The study highlighted significant disparities in vaccination coverage among children in Sub-Saharan Africa, influenced by sociodemographic and economic factors. Investing in maternal education, improving economic conditions, and strengthening healthcare infrastructure are essential measures to reduce these inequalities.

## Introduction

Vaccination is widely recognized as one of the most effective public health interventions for preventing infectious diseases and reducing child mortality [1]. The World Health Organization (WHO) estimates that immunization prevents between 2 to 3 million deaths annually worldwide [2]. However, vaccine coverage remains unequal, particularly in low- and middle-income countries, where social, economic, and cultural barriers can limit access to vaccines [3, 4].

Sub-Saharan Africa (SSA) is one of the regions facing the greatest challenges in terms of vaccine coverage. Disparities exist both between and within countries, with significant differences associated with factors such as maternal education level, economic status, geographical location (urban versus rural), and access to information [5, 6]. These disparities result in heightened vulnerability among underserved populations, exacerbating health inequities. Previous studies have demonstrated that maternal education is one of the strongest determinants of child vaccination coverage, with more educated mothers being more likely to adhere to their children’s vaccination schedules [7, 8]. Moreover, families with higher economic status have better access to healthcare services and are more likely to complete vaccination schedules [9]. Urban healthcare infrastructure also facilitates higher vaccination coverage compared to rural areas, where service availability is limited [4].

In the context of SSA, where cultural and religious diversity is significant, religious affiliation has also been identified as a factor influencing vaccination acceptance [8, 10]. For example, studies indicate that children from Muslim families have lower rates of full vaccination compared to those from Christian families in some countries [7]. This study aims to explore disparities in vaccine coverage among children aged 12 to 23 months in 10 SSA countries, identifying the key social and economic determinants influencing these disparities.

## Methods

### Study design and data Sources

This study utilized data from the Demographic and Health Surveys (DHS) and Multiple Indicator Cluster Surveys (MICS) of ten SSA countries. The most recent secondary datasets from Ghana, Nigeria, Liberia, Guinea, Gambia, Ethiopia, Côte d’Ivoire, Benin, Burkina Faso, and Cameroon were analyzed (Figure 1). DHS and MICS are nationally representative surveys that collect data on key health indicators, such as maternal and child health, mortality, morbidity, family planning services use, and vaccination coverage [11]. These datasets are accessible upon request and approval via the following link: https://www.dhsprogram.com/Data.

**Figure 1.**
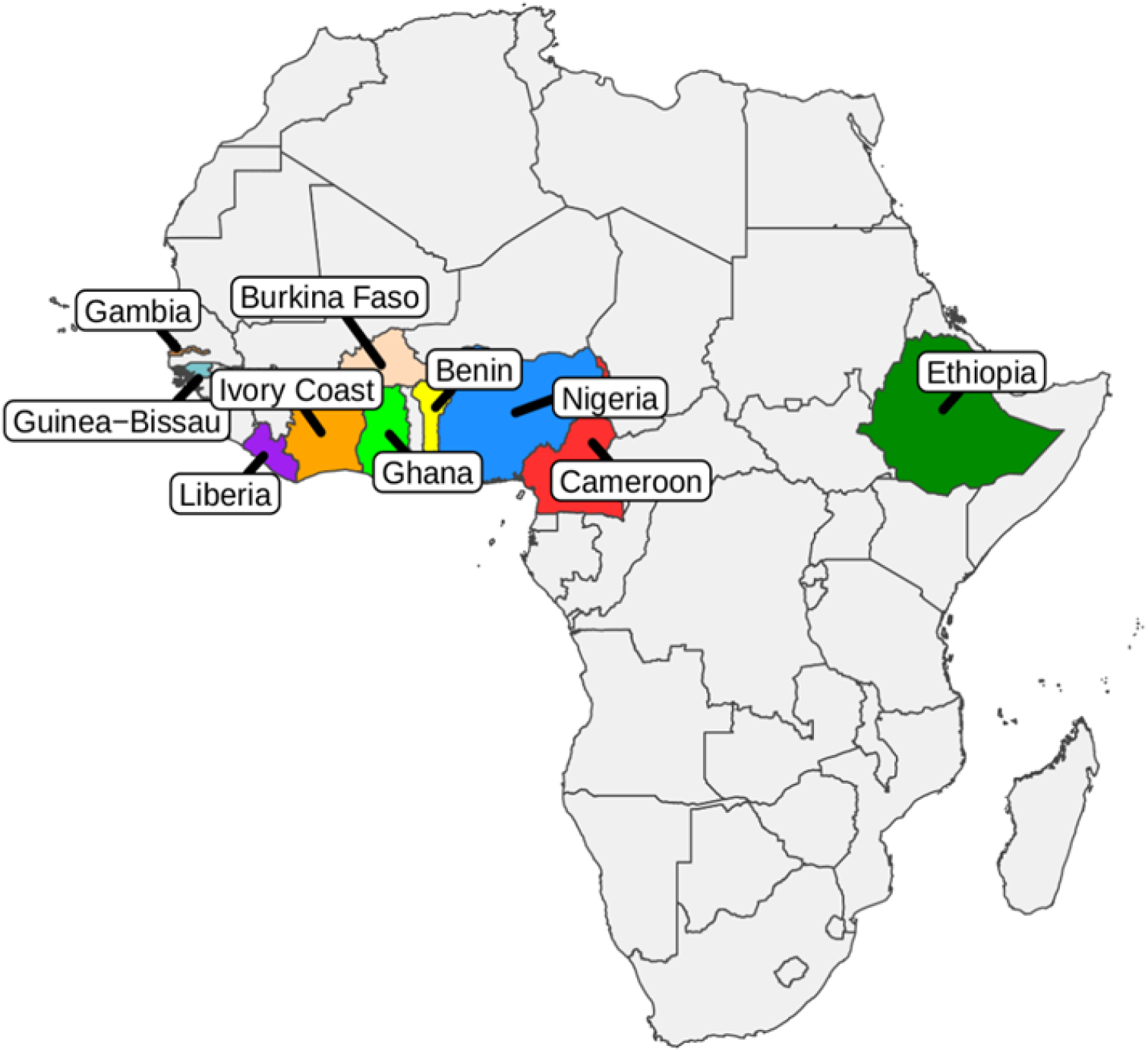
Map of African countries participating in the study on vaccination coverage among children aged 12 to 23 months in 10 Sub-Saharan African countries.

### Study Variables

The dependent variable was full vaccination coverage, defined as a child receiving one dose of BCG vaccine, three doses of the polio vaccine, three doses of the pentavalent vaccine (DPT), and one dose of the measles vaccine [12, 13].

The independent variables included religion (Christian, Muslim, and others), parental education level (none, primary, secondary, and higher), mother’s marital status (single, married/cohabiting, divorced/separated, and widowed), mother’s occupation (unemployed, informal sector, formal sector, and self-employed), access to prenatal care (none, limited, and adequate), type of residence (urban or rural), proximity to healthcare facilities (<1 km, 1–5 km, >5 km), access to information (presence of TV and/or radio), and family economic status (stratified into wealth quintiles: very poor, poor, middle, rich, very rich).

### Statistical analysis

Descriptive analysis was initially performed, including frequencies, means, and 95% confidence intervals for each vaccination indicator by country. Associations between full vaccination coverage and independent variables were tested using the chi-square or Fisher’s exact test. Logistic regression models were then applied to identify factors associated with full vaccination coverage, adjusting for potential confounding variables. Data analysis was conducted using R software version 4.3.2 (https://www.r-project.org/).

## Results

### Vaccination coverage and disparities

Full vaccination coverage among children aged 12 to 23 months in ten Sub-Saharan African countries exhibited wide variation, highlighting significant disparities across the region. Gambia recorded the highest coverage rate at 86.4% (95% CI: 84.5–88), while Guinea had the lowest, with only 21.2% (95% CI: 19.1–23.5). In Ghana, BCG vaccine coverage was 95.1% (95% CI: 93.9–96), and DPT1 coverage reached 96.9% (95% CI: 96.1–97.7). In contrast, Nigeria showed low coverage rates for DPT3 and Polio 3 vaccines, with 50.1% (95% CI: 48.8–51.4) and 47.2% (95% CI: 45.9–48.7), respectively (Table 1). Furthermore, Guinea stood out negatively with low measles vaccine coverage, achieving only 37.2% (95% CI: 34.7–39.8) **(Figures 1 and 2)**.

**Table 1:** Vaccination coverage in children aged 12 to 23 months in 10 Sub-Saharan African countries (2017-2022)

vacinal por país
| Immunization | Ghana | Nigeria | Liberia | Guinea | Gambia | Ethiopia | Côte d'Ivoire | Benin | Burkina-Faso | Cameroon |
| --- | --- | --- | --- | --- | --- | --- | --- | --- | --- | --- |
|  | (N = 1896) | (N = 5982) | (N = 1021) | (N = 1378) | (N = 1527) | (N = 1688) | (N = 1752) | (N = 2327) | (N = 2257) | (N = 1688) |
|  | 2022<br>% (CI 95%) | 2018<br>% (CI 95%) | 2019-20<br>% (CI 95%) | 2018<br>% (CI 95%) | 2019-20<br>% (CI 95%) | 2019<br>% (CI 95%) | 2021<br>% (CI 95%) | 2017-18<br>% (CI 95%) | 2021<br>% (CI 95%) | 2018<br>% (CI 95%) |
| BCG | 95.1 (93.9; 96.0) | 66.4 (65.2; 67.6) | 91.4 (89.4; 92.9) | 71.8 (69.4; 74.2) | 98.8 (98.1; 99.3) | 86.9 (85.2; 88.5) | 86.8 (85.1; 88.3) | 88.0 (86.6; 89.2) | 97.2 (96.4; 97.8) | 86.9 (85.2; 88.5) |
| DPT 1 | 96.9 (96.1; 97.7) | 64.4 (63.2; 65.6) | 91.5 (89.5; 93.1) | 60.4 (57.8; 63.0) | 98.4 (97.5; 98.9) | 82.6 (80.7; 84.4) | 73.8 (71.7; 75.8) | 84.1 (82.5; 85.5) | 95.4 (94.4; 96.2) | 82.6 (80.7; 84.4) |
| DPT 2 | 94.6 (93.4; 95.5) | 57.3 (56.0; 58.5) | 83.2 (80.7; 85.4) | 47.6 (44.9; 50.3) | 96.7 (95.6; 97.5) | 78.9 (76.8; 80.8) | 66.1 (63.8; 68.3) | 78.6 (76.8; 80.2) | 93.0 (91.9; 94.0) | 78.9 (76.8; 80.8) |
| DPT 3 | 89.3 (87.8; 90.6) | 50.1 (48.8; 51.4) | 68.8 (65.8; 71.6) | 38.0 (35.6; 40.7) | 94.6 (93.3; 95.6) | 72.9 (70.7; 75.0) | 57.5 (55.1; 59.8) | 73.8 (71.9; 75.6) | 89.2 (87.8; 90.4) | 72.9 (70.7; 75.0) |
| Polio 0 | 74.4 (72.3; 76.3) | 53.6 (52.4; 54.9) | 84.1 (81.7; 86.3) | 67.5 (64.9; 69.9) | 98.3 (97.5; 98.9) | 76.2 (74.1; 78.2) | 69.7 (67.5; 71.1) | 86.3 (84.8; 87.6) | 93.6 (92.5; 94.5) | 76.2 (74.1; 78.2) |
| Polio 1 | 95.1 (94.1; 96.1) | 73.1 (71.9; 74.2) | 87.6 (85.3; 89.5) | 62.6 (59.9; 65.1) | 97.6 (96.6; 98.3) | 85.1 (83.3; 86.7) | 82.1 (80.2; 83.8) | 82.1 (80.5; 83.6) | 92.8 (91.6; 93.8) | 85.1 (83.3; 86.7) |
| Polio 2 | 91.3 (89.9; 92.5) | 66.7 (65.5; 67.9) | 78.8 (76.2; 81.3) | 50.1 (47.5; 52.8) | 96.0 (94.9; 96.9) | 79.5 (77.5; 81.4) | 75.1(73.0; 77.1) | 77.5 (75.8; 79.2) | 89.9 (88.5; 91.1) | 79.5 (77.5; 81.4) |
| Polio 3 | 83.9 (82.2; 85.5) | 47.2 (45.9; 48.7) | 62.9 (59.8; 65.8) | 37.7 (35.1; 40.3) | 92.9(91.4; 94.1) | 69.0 (66.7; 71.1) | 63.2(60.9; 65.4) | 66.1(64.2; 68.1) | 82.0(80.4; 83.6) | 69.0(66.7; 71.1) |
| Sarampo 1 | 86.7 (85.1; 88.2) | 53.1 (51.8; 54.4) | 71.5 (68.6; 74.2) | 37.2 (34.7; 39.8) | 92.1 (90.6; 93.4) | 65.5 (63.1; 67.8) | 65.0 (62.7; 67.2) | 68.0 (66.1; 69.9) | 85.8 (84.3; 87.2) | 65.5 (63.2; 67.8) |
| Coverage |  |  |  |  |  |  |  |  |  |  |
| Complete | 74.6 (72.5; 76.5) | 30.5 (29.4; 31.7) | 49.8 (46.6; 52.9) | 21.2 (19.1; 23.5) | 86.4 (84.5; 88.0) | 53.1 (50.7; 55.5) | 38.8 (36.5; 41.1) | 56.5 (54.4; 58.5) | 72.4 (70.5; 74.3) | 53.1 (50.7; 55.5) |
| Incomplete | 25.4 (23.5; 27.5) | 69.5 (68.3; 70.6) | 50.2 (47.1; 53.4) | 78.8 (76.5; 80.9) | 13.6 (12.0; 15.5) | 46.9 (44.5; 49.3) | 61.2 (58.9; 63.5) | 43.5 (41.5; 45.6) | 27.6 (25.7; 29.5) | 46.9 (44.5; 49.3) |

**Figure 2.**
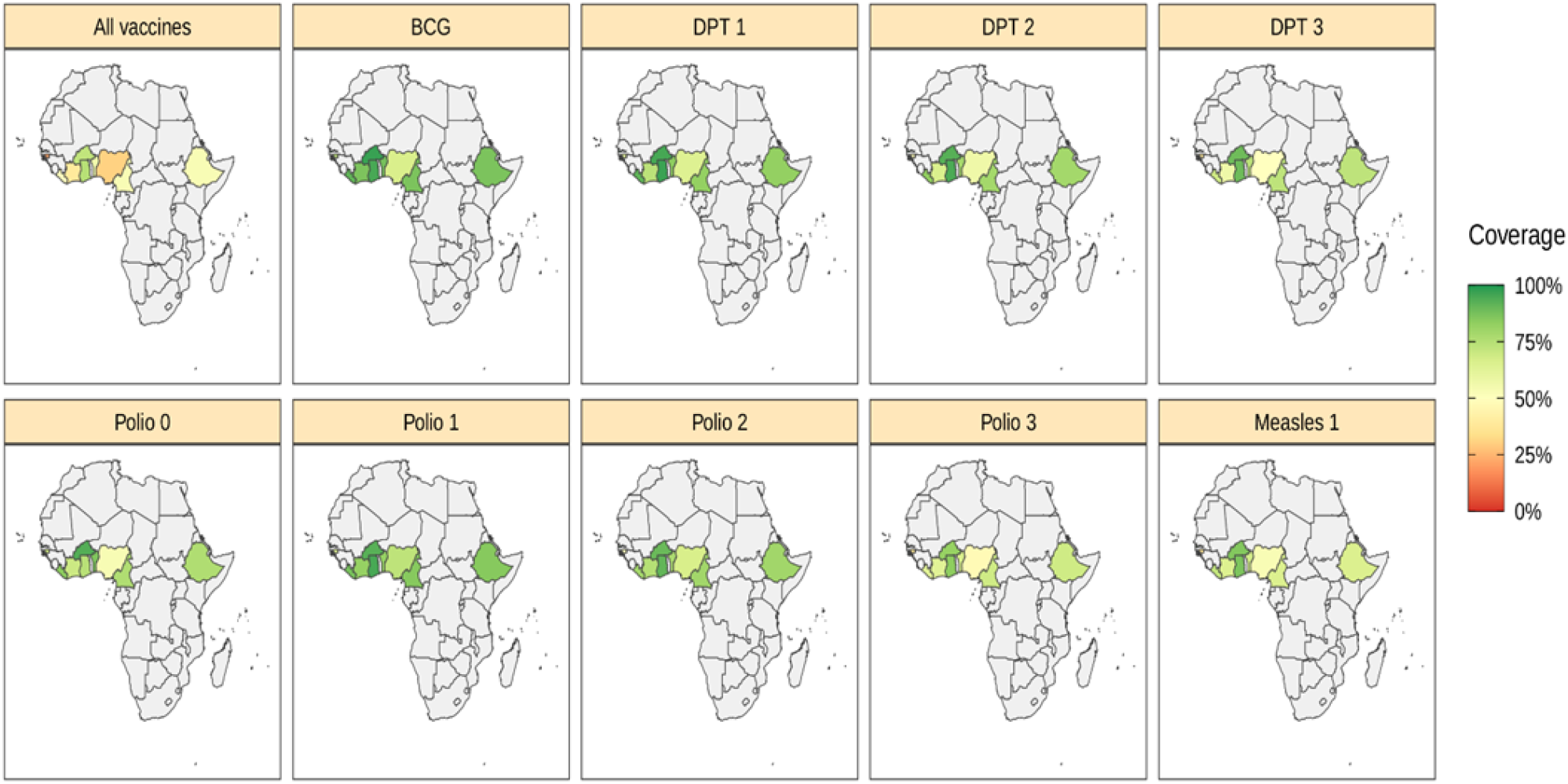
Percentage of vaccination coverage among children aged 12 to 23 months for different vaccines in SSA (2017–2022).

### Vaccination coverage

Various sociodemographic and economic factors were associated with full vaccination coverage, as presented in Table 2. Religion significantly influenced vaccination rates, with children from Christian families in countries such as Ghana, Nigeria, and Cameroon exhibiting higher coverage compared to those from other religions (p < 0.001). Similarly, urban residence was consistently associated with better vaccination rates in countries like Burkina Faso, Ghana, and Cameroon (p < 0.01). Maternal education also emerged as a critical factor, with children of mothers with higher education achieving higher vaccination coverage in Ghana, Benin, and Nigeria (p < 0.001). Family economic status was another significant determinant, with children from wealthier families showing higher vaccination coverage than those from poorer families, as observed in Côte d’Ivoire (68.0% versus 25.2%; p < 0.001). Other factors, such as proximity to healthcare facilities and access to information, also influenced vaccination rates.

**Table 2.**
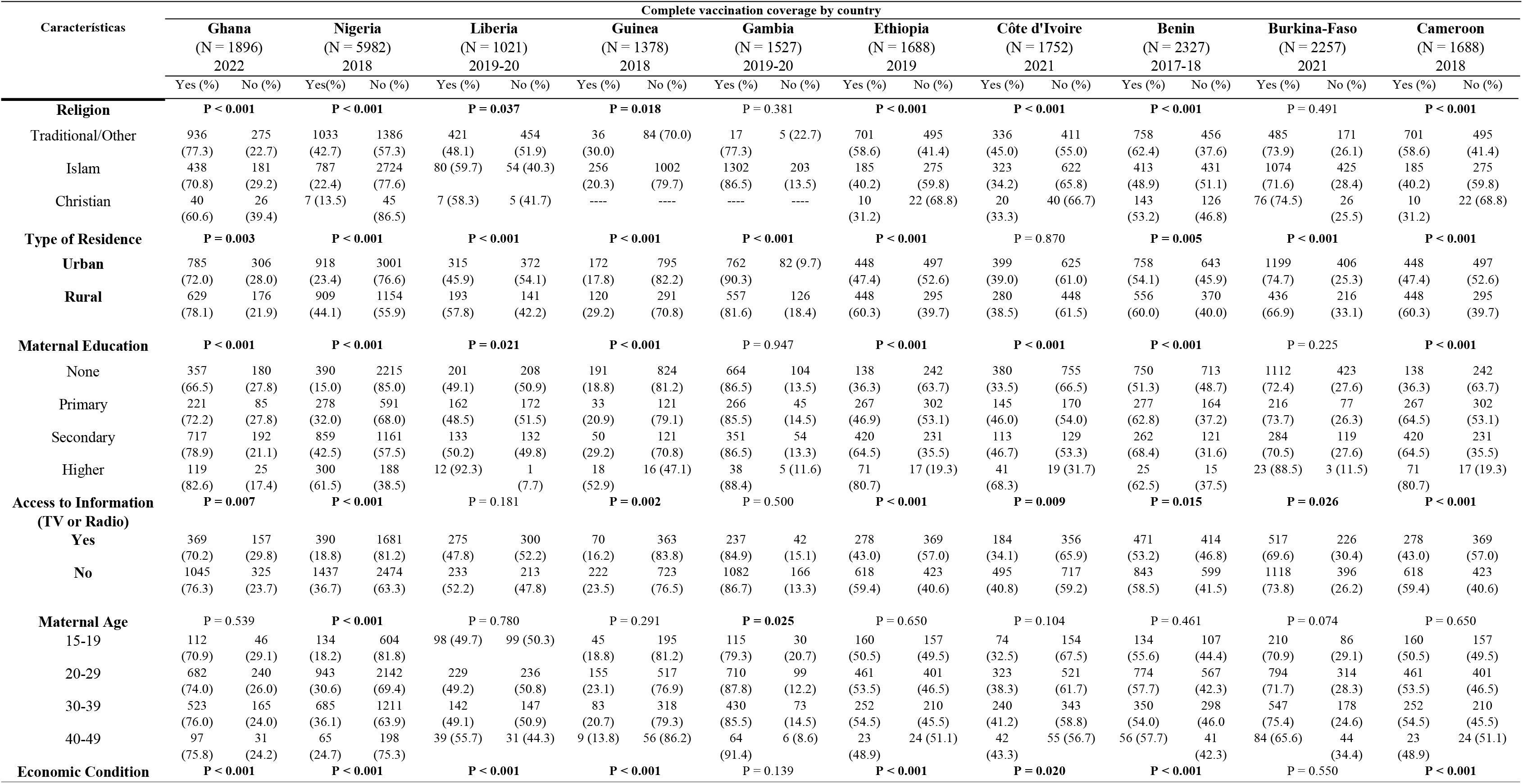
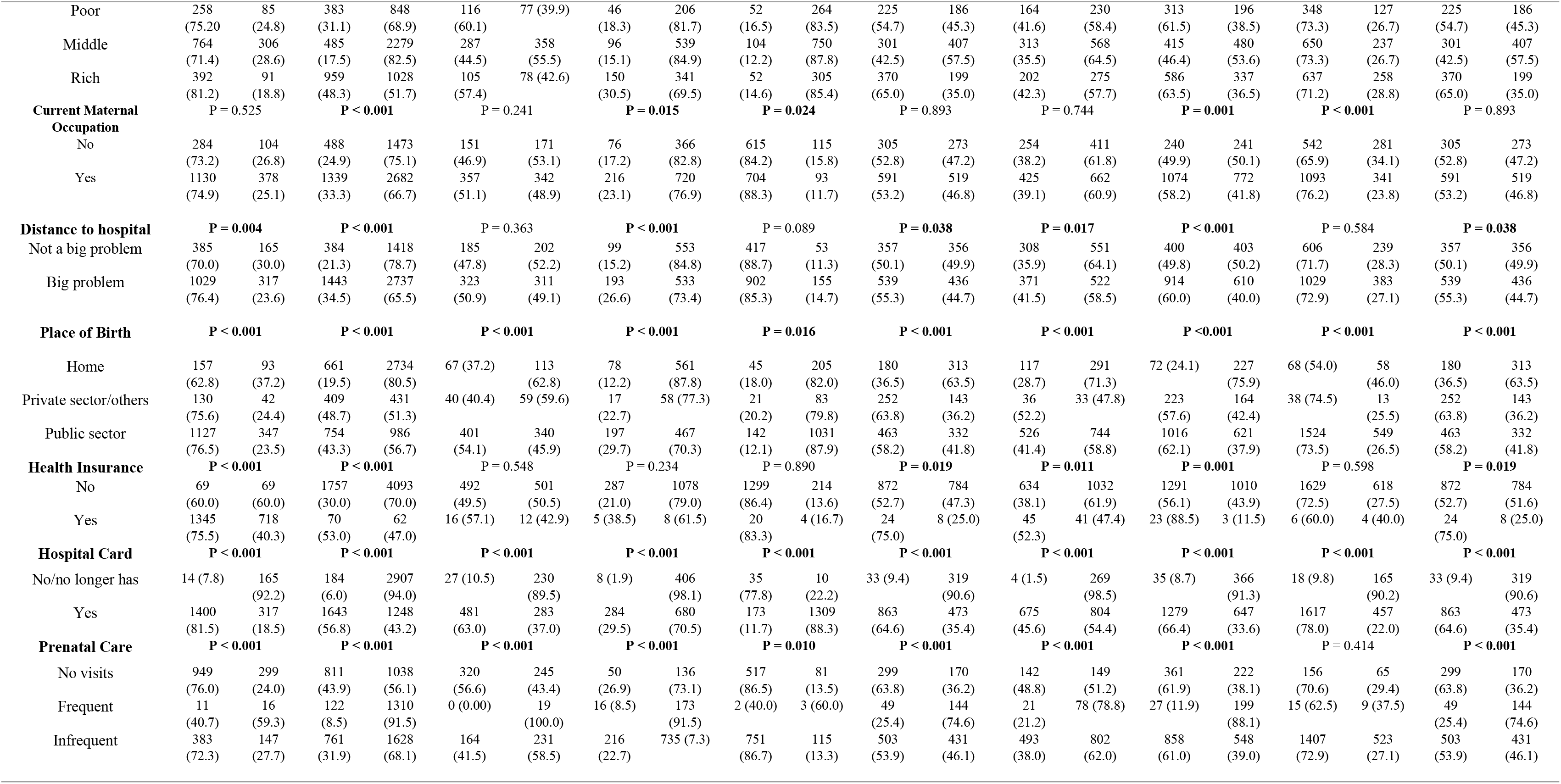
Inequities in complete vaccination coverage by country, related to socioeconomic, geographic, maternal and child health characteristics (2017-2022)

### Logistic models of factors associated with vaccination coverage

Tables 3, 4, and 5 present crude and adjusted logistic regression models, highlighting maternal higher education as a key factor for full vaccination coverage. In Ghana, children of mothers with higher education were 3.6 times more likely to be fully vaccinated compared to those whose mothers had no education (AOR: 3.60; 95% CI: 1.93–6.72). Similar results were observed in Côte d’Ivoire, Cameroon, Nigeria, Guinea, and Ethiopia, with AORs ranging from 2.21 to 4.97.

**Table 3.** Crude logistic regression models of the associations between socioeconomic, geographic, maternal and child health characteristics and vaccination coverage (2017-2022)

|  | Ghana | Nigeria | Liberia | Guinea | Gambia | Ethiopia | Côte d'Ivoire | Benin | Burkina-Faso | Cameroon |
| --- | --- | --- | --- | --- | --- | --- | --- | --- | --- | --- |
|  | COR | COR | COR | COR | COR | COR | COR | COR | COR | COR |
|  | (CI 95%) | (CI 95%) | (CI 95%) | (CI 95%) | (CI 95%) | (CI 95%) | (CI 95%) | (CI 95%) | (CI 95%) | (CI 95%) |
| **Religion** |  |  |  |  |  |  |  |  |  |  |
| Traditional/Other | Ref | Ref | Ref |  |  | Ref | Ref | Ref | Ref | Ref |
| Islam | 1.57 (0.93; 2.65) | 1.86 (0.83; 4.13) | 1.06 (0.32; 3.51) | Ref | Ref | 1.48 (0.68; 3.20) | 1.04 (0.60; 1.81) | 0.84 (0.64; 1.11) | 0.86 (0.55; 1.37) | 1.48 (0.68; 3.20) |
| Christian | 2.21 (1.33; 3.69) | 4.79 (2.15; 10.67) | 0.66 (0.21; 2.10) | 1.68 (1.11; 2.54) | 1.89 (0.69; 5.17) | 3.12 (1.46; 6.64) | 1.64 (0.94; 2.85) | 1.46 (1.12; 1.91) | 0.97 (0.60; 1.57) | 3.12 (1.46; 6.64) |
| **Type of Residence** |  |  |  |  |  |  |  |  |  |  |
| Urban | Ref | Ref | Ref | Ref | Ref | Ref | Ref | Ref | Ref | Ref |
| Rural | 0.72 (0.58; 0.89) | 0.39 (0.35; 0.44) | 0.62 (0.48; 0.81) | 0.52 (0.40; 0.69) | 0.48 (0.35; 0.64) | 0.59 (0.49; 0.72) | 1.02 (0.84; 1.24) | 0.78 (0.66; 0.93) | 1.46 (1.20; 1.78) | 0.59 (0.49; 0.72) |
| **Maternal Education** |  |  |  |  |  |  |  |  |  |  |
| None | Ref | Ref | Ref | Ref | Ref | Ref | Ref | Ref | Ref | Ref |
| Primary | 1.31 (0.96; 1.78) | 2.67 (2.23; 3.19) | 0.97 (0.73; 1.30) | 1.14 (0.75; 1.72) | 1.08 (0.74; 1.58) | 1.55 (1.19; 2.02) | 1.69 (1.32; 2.18) | 1.61 (1.29; 2.00) | 1.07 (0.80; 1.42) | 3.19 (2.45; 4.15) |
| Secondary | 1.88 (1.48; 2.39) | 4.20 (3.66; 4.83) | 1.04 (0.77; 1.42) | 1.78 (1.24; 2.57) | 0.98 (0.69; 1.40) | 3.19 (2.45; 4.15) | 1.74 (1.31; 2.31) | 2.06 (1.62; 2.61) | 0.91 (0.71; 1.16) | 3.19 (2.45; 4.15) |
| Higher | 2.40 (1.50; 3.83) | 9.06 (7.33; 11.20) | 12.4 (1.60; 96.24) | 4.85 (2.43; 9.69) | 0.84 (0.32; 2.18) | 7.32 (4.2; 12.9) | 1.02 (0.84; 1.24) | 1.58 (0.83; 3.03) | 2.92 (0.87; 9.76) | 7.32 (4.15; 12.9) |
| **Access to Information (TV or Radio)** |  |  |  |  |  |  |  |  |  |  |
| Yes | Ref | Ref | Ref | Ref | Ref | Ref | Ref | Ref | Ref | Ref |
| No | 0.73 (0.58; 0.91) | 0.40 (0.35; 0.45) | 0.84 (0.65; 1.07) | 0.63 (0.47; 0.85) | 1.16 (0.80; 1.67) | 0.52 (0.42; 0.63) | 0.75 (0.61; 0.93) | 0.81 (0.68; 0.96) | 0.81 (0.67; 0.98) | 0.52 (0.42; 0.63) |
| **Maternal Age** |  |  |  |  |  |  |  |  |  |  |
| 15-19 | Ref | Ref | Ref | Ref | Ref | Ref | Ref | Ref | Ref | Ref |
| 20-29 | 1.17 (0.80; 1.70) | 1.98 (1.62; 2.43) | 0.98 (0.70; 1.37) | 1.30 (0.90; 1.88) | 0.53 (0.34; 0.84) | 1.13 (0.87; 1.46) | 1.29 (0.95; 1.76) | 1.09 (0.83; 1.44) | 1.04 (0.78; 1.37) | 1.13 (0.87; 1.46) |
| 30-39 | 1.30 (0.89; 1.91) | 2.55 (2.07; 3.14) | 0.98 (0.68; 1.40) | 1.13 (0.75; 1.69) | 0.65 (0.41; 1.04) | 1.18 (0.88; 1.57) | 1.46 (1.05; 2.01) | 0.94 (0.70; 1.26) | 1.26 (0.93; 1.70) | 1.18 (0.88; 1.57) |
| 40-49 | 1.29 (0.76; 2.18) | 1.48 (1.06; 2.07) | 1.27 (0.73; 2.20) | 0.70 (0.32; 1.51) | 0.36 (0.14; 0.91) | 0.94 (0.51; 1.74) | 1.59 (0.98; 2.59) | 1.09 (0.68; 1.76) | 0.78 (0.50; 1.22) | 0.52 (0.42; 0.63) |
| **Economic Condition** |  |  |  |  |  |  |  |  |  |  |
| Poor | Ref | Ref | Ref | Ref | Ref | Ref | Ref | Ref | Ref | Ref |
| Middle | 1.22 (0.92; 1.61) | 2.12 (1.82; 2.48) | 1.88 (1.35; 2.61) | 1.25 (0.85; 1.85) | 1.42 (0.99; 2.04) | 1.64 (1.28; 2.09) | 1.29 (1.01; 1.65) | 1.85(1.48; 2.31 | 1.00 (0.78; 1.29) | 1.64 (1.28; 2.09) |
| Rich | 1.73 (1.33; 2.25) | 4.38 (3.84; 5.00) | 1.68 (1.21; 2.34) | 2.47 (1.85; 3.30) | 1.23 (0.86; 1.76) | 2.51 (2.00; 3.16) | 1.33 (1.06; 1.67) | 2.01 (1.67; 2.43) | 0.90 (0.73; 1.11) | 2.51 (2.00; 3.16) |
| **Distance to hospital** |  |  |  |  |  |  |  |  |  |  |
| Not a big problem | Ref | Ref | Ref | Ref | Ref | Ref | Ref | Ref | Ref | Ref |
| Big problem | 0.72 (0.58; 0.90) | 0.51 (0.45; 0.58) | 0.88 (0.68; 1.14) | 0.49 (0.38; 0.65) | 0.74 (0.53; 1.03) | 0.81 (0.67; 0.98) | 0.79 (0.65; 0.95) | 0.66 (0.56; 0.79) | 0.94 (0.78; 1.14) | 0.81 (0.67; 0.98) |
| **Place of Birth** |  |  |  |  |  |  |  |  |  |  |
| Home | Ref | Ref | Ref | Ref | Ref | Ref | Ref | Ref | Ref | Ref |
| Private sector/others | 1.83 (1.19; 2.82) | 3.93 (3.35; 4.60) | 1.14 (0.69; 1.89) | 2.11 (1.17; 3.80) | 1.15 (0.65; 2.05) | 3.06 (2.33; 4.04) | 2.71 (1.62; 4.56) | 4.29 (3.07; 5.98) | 2.49 (1.21; 5.13) | 3.06 (2.33; 4.04) |
| Public sector | 1.92 (1.45; 2.550) | 3.16 (2.78; 3.59) | 1.99 (1.42; 2.78) | 3.03 (2.27; 4.05) | 0.63 (0.43; 0.91) | 2.43 (1.92; 3.06) | 1.76 (1.38; 2.24) | 5.16 (3.89; 6.85) | 2.37 (1.65; 3.41) | 2.43 (1.92; 3.06) |
| **Health Insurance** |  |  |  |  |  |  |  |  |  |  |
| No | Ref | Ref | Ref | Ref | Ref | Ref | Ref | Ref | Ref | Ref |
| Yes | 2.06 (1.39; 3.03) | 2.63 (1.86; 3.72) | 1.36 (0.64; 2.90) | 2.35 (0.76; 7.23) | 1.27 (0.43; 3.76) | 2.70 (1.20; 6.04) | 1.79 (1.16; 2.76) | 6.0 (1.80; 20.03) | 0.57 (0.16; 2.02) | 2.70 (1.20; 6.04) |
| **Hospital Card** |  |  |  |  |  |  |  |  |  |  |
| No/no longer has | Ref | Ref | Ref | Ref | Ref | Ref | Ref | Ref | Ref | Ref |
| Yes | 52 (29.76; 91.04) | 20.8 (17.61; 24.56) | 14.48 (9.47; 22.14) | 21.2 (10.39; 43.3) | 0.04 (0.02; 0.08) | 17.64 (12.; 25.7) | 56.5 (20.9; 152) | 20.7 (14.4; 29.6) | 32.4 (19.7; 53.3) | 17.6(12.1 \| 25.7) |
| **Prenatal Care** |  |  |  |  |  |  |  |  |  |  |
| No visits | Ref | Ref | Ref | Ref | Ref | Ref | Ref | Ref | Ref | Ref |
| Frequent | 0.22 (0.10; 0.47) | 0.12 (0.10; 0.15) | 0.01 (0.00; 2.14) | 0.25 (0.14; 0.46) | 9.6 (1.58; 58.1) | 0.19 (0.13; 0.28) | 0.28 (0.17; 0.48) | 0.08 (0.05; 0.13) | 0.69 (0.29; 1.67) | 0.19 (0.13; 0.28) |
| Infrequent | 0.82 (0.65; 1.03) | 0.60 (0.53; 0.68) | 0.54 (0.42; 0.71) | 0.80 (0.56; 1.14) | 0.98 (0.72; 1.33) | 0.66 (0.53; 0.83) | 0.64 (0.50; 0.83) | 0.96 (0.79; 1.17) | 1.12 (0.83; 1.52) | 0.66 (0.53; 0.83) |

**Table 4:** Adjusted logistic regression models of associations between socioeconomic, geographic, maternal and child health characteristics and vaccination coverage (DHS and MIS, 2017-2022)

|  |  |  |  |  |  |  |  |  |  |  |
| --- | --- | --- | --- | --- | --- | --- | --- | --- | --- | --- |
|  | Ghana | Nigeria | Liberia | Guinea | Gambia | Ethiopia | Côte d'Ivoire | Benin | Burkina-Faso | Cameroon |
|  | AOR | AOR | AOR | AOR | AOR | AOR | AOR | AOR | AOR | AOR |
|  | (CI 95%) | (CI 95%) | (CI 95%) | (CI 95%) | (CI 95%) | (CI 95%) | (CI 95%) | (CI 95%) | (CI 95%) | (CI 95%) |
| **Religion** |  |  |  |  |  |  |  |  |  |  |
| Traditional/Other | Ref | Ref | Ref |  |  | Ref | Ref | Ref | Ref | Ref |
| Islam | 1.30 (0.70; 2.43) | 2.06 (0.79; 5.39) | 1.00 (0.26; 3.91) | Ref | Ref | 1.51 (0.62; 3.69) | 0.92 (0.50; 1.70) | 1.12 (0.80; 1.55) | 0.82 (0.47; 1.42) | 1.51 (0.62; 3.69) |
| Christian | 1.05 (0.56; 1.98) | 2.29 (0.88; 5.96) | 0.66 (0.18; 2.46) | 1.33 (0.83; 2.13) | 1.18 (0.37; 3.72) | 1.94 (0.81; 4.66) | 1.12 (0.61; 2.07) | 1.21 (0.88; 1.65) | 0.88 (0.50; 1.55) | 1.94 (0.81; 4.66) |
| **Type of Residence** |  |  |  |  |  |  |  |  |  |  |
| Urban | Ref | Ref | Ref | Ref | Ref | Ref | Ref | Ref | Ref | Ref |
| Rural | 0.95 (0.70; 1.29) | 0.89 (0.76; 1.06) | 0.81 (0.57; 1.16) | 1.34 (0.85; 2.10) | 0.34 (0.22; 0.53) | 1.11 (0.82; 1.51) | 1.57 (1.18; 2.08) | 1.05 (0.84; 1.20) | 1.58 (1.17; 2.13) | 1.11 (0.82; 1.51) |
| **Maternal Education** |  |  |  |  |  |  |  |  |  |  |
| None | Ref | Ref | Ref | Ref | Ref | Ref | Ref | Ref | Ref | Ref |
| Primary | 1.21 (0.83; 1.77) | 1.20 (0.95; 1.51) | 1.12 (0.79; 1.60) | 0.89 (0.57; 1.41) | 0.97 (0.63; 1.49) | 0.89 (0.63; 1.26) | 1.45 (1.08; 1.96) | 1.22 (0.94; 1.58) | 1.00 (0.72; 1.39) | 0.89 (0.63; 1.26) |
| Secondary | 1.54 (1.10; 2.16) | 1.36 (1.08; 1.70) | 1.17 (0.77; 1.78) | 1.00 (0.65; 1.56) | 0.91(0.60; 1.39) | 1.38 (0.93; 2.06) | 1.62 (1.14; 2.29) | 1.34 (0.99; 1.80) | 0.95 (0.70; 1.29) | 1.38 (0.93; 2.06) |
| Higher | 2.80 (1.38; 5.67) | 1.97 (1.42; 2.73) | 9.35 (0.98; 89.14) | 2.96 (1.25; 7.01) | 0.70(0.23; 2.15) | 2.81 (1.32; 5.95) | 4.43 (2.13; 9.22) | 1.15 (0.52; 2.53) | 3.79 (0.91; 15.85) | 2.81 (1.32; 5.95) |
| **Maternal Age** |  |  |  |  |  |  |  |  |  |  |
| 15-19 | Ref | Ref | Ref | Ref | Ref | Ref | Ref | Ref | Ref | Ref |
| 20-29 | 2.80 (1.38; 5.67) | 1.60 (1.25; 2.04) | 1.11 (0.75; 1.62) | 1.26 (0.83; 1.91) | 0.42 (0.26; 0.70) | 0.98 (0.72; 1.34) | 1.24 (0.88; 1.75) | 1.12 (0.81; 1.55) | 1.00 (0.72; 1.38) | 0.98 (0.72; 1.34) |
| 30-39 | 0.98 (0.61; 1.58) | 1.96 (1.51; 2.55) | 1.02 (0.66; 1.57) | 1.19 (0.75; 1.87) | 0.57 (0.34; 0.96) | 1.13 (0.80; 1.62) | 1.50 (1.04; 2.15) | 1.04 (0.73; 1.48) | 1.30 (0.91; 1.85) | 1.13 (0.80 (1.62) |
| 40-49 | 1.29 (0.67; 2.47) | 1.38 (0.90; 2.10) | 1.43; 0.74 (2.76) | 0.86; 0.37 (2.01) | 0.31; 0.11 (0.84) | 1.00; 0.49 (2.05) | 1.62; 0.94 (2.79) | 1.33; 0.75 (2.34) | 0.80 (0.48; 1.33) | 1.00 (0.49; 2.05) |
| **Economic Condition** |  |  |  |  |  |  |  |  |  |  |
| Poor | Ref | Ref | Ref | Ref | Ref | Ref | Ref | Ref | Ref | Ref |
| Middle | 0.90 (0.63; 1.30) | 1.01 (0.83; 1.24) | 1.84 (1.23; 2.73) | 1.16 (0.76; 1.76) | 0.73 (0.45; 1.17) | 1.26 (0.92; 1.74) | 1.29 (0.96; 1.73) | 1.30 (1.01; 1.69) | 1.11 (0.83; 1.49) | 1.26 (0.92; 1.74) |
| Rich | 1.16 (0.77; 1.75) | 1.42 (1.15; 1.77) | 1.61 (0.98; 2.64) | 2.10 (1.30; 3.38) | 0.58 (0.35; 0.98) | 1.54 (1.05; 2.26) | 1.14 (0.80; 1.62) | 1.17 (0.90; 1.52) | 1.26 (0.93; 1.72) | 1.26 (0.92; 1.74) |
| **Distance to hospital** |  |  |  |  |  |  |  |  |  |  |
| Not a big problem | Ref | Ref | Ref | Ref | Ref | Ref | Ref | Ref | Ref | Ref |
| Big problem | 0.99 (0.75; 1.31) | 0.81 (0.68; 0.96) | 1.20 (0.88; 1.63) | 0.74 (0.54; 1.01) | 0.82 (0.55; 1.22) | 1.24 (0.97; 1.58) | 0.94 (0.75; 1.18) | 0.93 (0.75; 1.14) | 1.00 (0.79; 1.27) | 1.24 (0.97; 1.58) |
| **Place of Birth** |  |  |  |  |  |  |  |  |  |  |
| Home | Ref | Ref | Ref | Ref | Ref | Ref | Ref | Ref | Ref | Ref |
| Private sector/others | 1.17 (0.68; 2.01) | 1.06 (0.84; 1.34) | 0.89 (0.49; 1.61) | 1.04 (0.53; 2.06) | 0.89 (0.45; 1.76) | 1.47 (1.01; 2.16) | 2.00 (1.08; 3.70) | 1.28 (0.82; 1.98) | 2.90 (1.24; 6.79) | 1.47 (1.01; 2.16) |
| Public sector | 1.51 (1.05; 2.16) | 1.18 (0.99; 1.41) | 1.46 (0.99; 2.16) | 1.77 (1.26; 2.49) | 0.50 (0.33; 0.76) | 1.24 (0.89; 1.71) | 1.63 (1.23; 2.17) | 1.56 (1.07; 2.27) | 2.72 (1.81; 4.11) | 1.24 (0.89; 1.71) |
| **Health Insurance** |  |  |  |  |  |  |  |  |  |  |
| No | Ref | Ref | Ref | Ref | Ref | Ref | Ref | Ref | Ref | Ref |
| Yes | 1.39 (0.86; 2.25) | 1.49 (0.92; 2.39) | 0.84 (0.34; 2.08) | 0.94 (0.27; 3.26) | 1.53 (0.48; 4.82) | 1.60 (0.61; 4.24) | 0.77 (0.46; 1.31) | 4.1 (1.11; 15.2) | 0.5(0.09; 2.41) | 1.60 (0.61; 4.24) |
| **Hospital Card** |  |  |  |  |  |  |  |  |  |  |
| No/no longer has | Ref | Ref | Ref | Ref | Ref | Ref | Ref | Ref | Ref | Ref |
| Yes | 51.4(28.8; 91.63) | 14.7 (12; 17.6) | 14.7 (9.50; 22.82) | 20.9(9.8; 44.7) | 0.03 (0.02; 0.08) | 15.6(10.5; 23.2) | 53.5 (19.7; 145) | 13.6 (9.28; 19.9) | 34.8(20.5; 59) | 15.62 (10.54; 23.16) |
| **Prenatal Care** |  |  |  |  |  |  |  |  |  |  |
| No visits | Ref | Ref |  | Ref | Ref | Ref | Ref | Ref | Ref | Ref |
| Frequent | 0.64 (0.22; 1.82) | 1.80 (1.39; 2.35) | — | 0.59 (0.28; 1.25) | 0.10 (0.01; 0.77) | 1.69 (1.04; 2.76) | 1.34 (0.71; 2.53) | 3.43 (2.03; 5.79) | 0.93 (0.30; 2.87) | 1.69 (1.04; 2.76) |
| Infrequent | 0.91 (0.69; 1.20) | 1.87 (1.46; 2.38) | — | 0.57 (0.29; 1.11) | 0.10 (0.01; 0.77) | 1.50 (0.97; 2.32) | 0.97 (0.55; 1.72) | 3.58 (2.19; 5.85) | 1.15 (0.38; 3.43) | 1.50 (0.97; 2.32) |

**Table 4aa.**
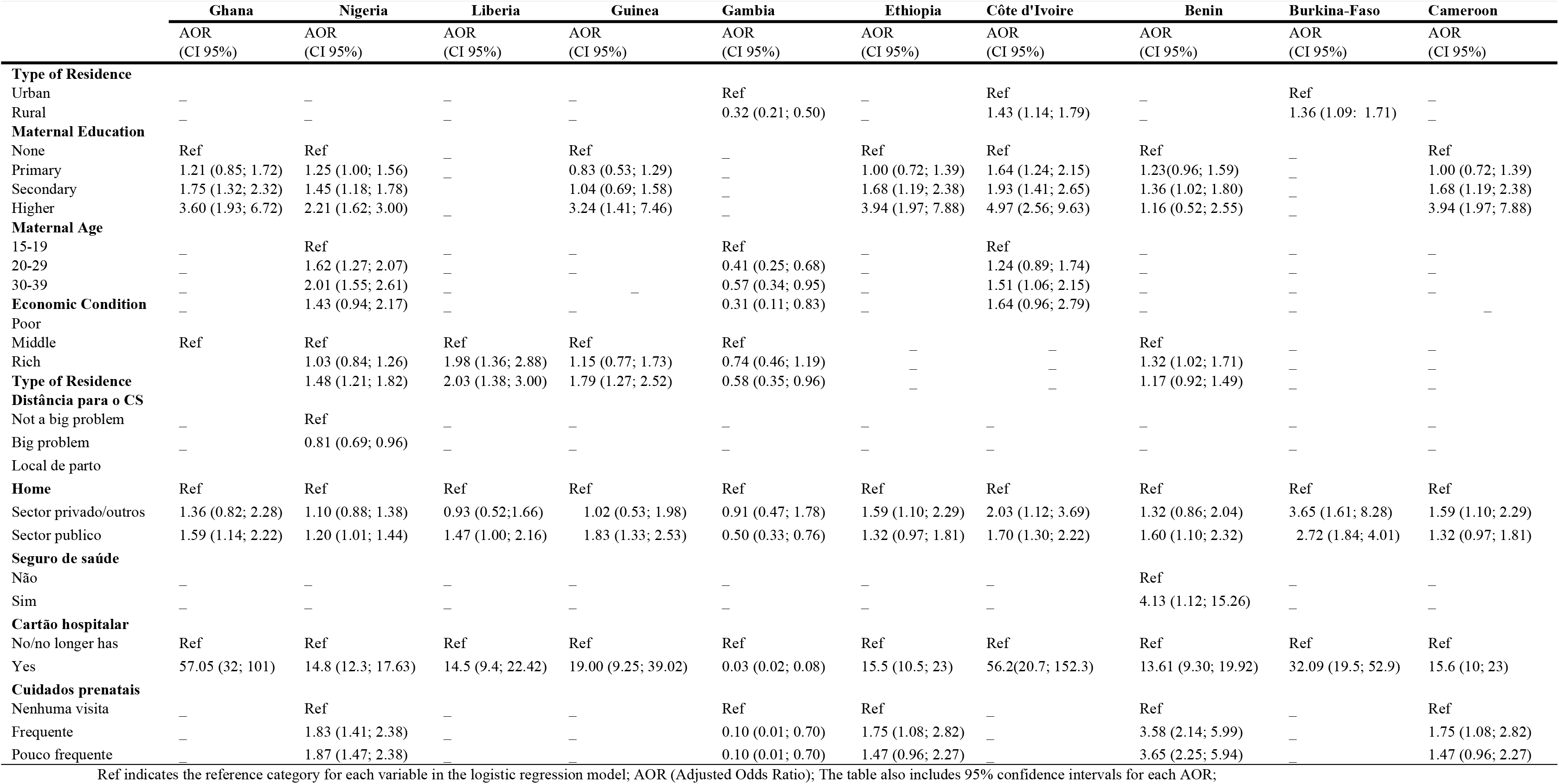
Adjusted logistic regression models analyzing the association between socio-demographic characteristics and complete vaccination coverage in children from SSA Countries (DHS & MIS, 2017-2022)

Residence also significantly influenced vaccination coverage. In Gambia, children in rural areas had only 32% of the odds of being fully vaccinated compared to those in urban areas (AOR: 0.32; 95% CI: 0.21–0.50). Conversely, in Burkina Faso and Côte d’Ivoire, children in rural areas showed higher vaccination coverage than those in urban settings. Family economic status was another critical determinant. Children from wealthy families in countries such as Nigeria, Liberia, and Guinea were significantly more likely to achieve full vaccination coverage compared to those from poorer families. Other factors included maternal age, with mothers aged 30–39 years in Cameroon and Côte d’Ivoire having higher odds of fully vaccinating their children compared to mothers aged 15–19 years. Additionally, children born in public institutions and those possessing a vaccination card were more likely to achieve full vaccination coverage.

## Discussion

This study revealed significant variation in full vaccination coverage rates across the analyzed countries. While Gambia (86.4%), Ghana (74.6%), and Burkina Faso (72.4%) demonstrated the highest rates, countries such as Nigeria, Liberia, Guinea, Côte d’Ivoire, and Benin reported coverage rates below 50%. These findings align with previous studies conducted in other contexts, such as Tanzania (71.1%) [6], Pakistan (74.1%) [14], Ethiopia (75.6%) [15], Southeast Asia (78.6%) [16] and Mozambique (71.8%) [17]. Additional research using DHS data found similar results for Ethiopia, with a rate of 58.4% [18], while more recent data from 2023 reported a lower rate of 39.09% [19]. In Nigeria, full vaccination coverage increased from 11% in 2013 to 18% in 2018 [20], rates still lower compared to the national coverage identified in this study (30.5%). In SSA, only about 60% of children aged 12–23 months receive complete immunization [21]. In comparison, India had a rate of 60.9% while China achieved 83.1% [22].

Although differences in full vaccination coverage rates may partially be attributed to variations in study designs and methodologies, the findings underscore inequalities in vaccination coverage both within SSAand in comparison, to other countries worldwide. These disparities can be attributed to differences in healthcare infrastructure quality, vaccination policies, awareness of immunization importance, and diverse sociocultural, geographic, and economic factors [13].

Bivariate analysis revealed significant associations between full vaccination coverage and factors such as religion, type of residence, maternal education, access to information, maternal age, economic status, maternal occupation, distance to healthcare facilities, place of birth, health insurance, possession of a vaccination card, and frequency of prenatal care visits. After adjusting for potential confounders, the determinants that remained significantly associated included residence, maternal education, maternal age, economic status, place of birth, possession of a vaccination card, and frequency of prenatal care visits.

In this study, children in urban areas generally showed lower vaccination coverage compared to those in rural areas, consistent with previous findings [12, 18, 19], although rural children in Gambia exhibited lower coverage. This discrepancy may be explained by limited awareness, difficulties accessing healthcare services, and low utilization of vaccination services among rural residents [23]. Interestingly, children of mothers with higher education showed greater vaccination coverage compared to those with uneducated mothers, a result supported by prior studies [17, 19, 24, 25]. These findings highlight the importance of education and awareness interventions to improve childhood vaccination coverage, especially among vulnerable populations. Regarding maternal age, results were mixed. In Gambia, children of adolescent mothers showed lower vaccination coverage compared to those of non-adolescent mothers, consistent with other studies [25]. This may be due to adolescent mothers having less contact with healthcare services, where they would receive guidance on childhood vaccination, thereby improving their children’s immunization coverage.

Concerning the place of birth, children born in public health institutions exhibited higher vaccination coverage in nearly all countries included in this study compared to those born outside such institutions. This finding is consistent with other research [12, 15, 18, 19, 23, 26, 27]. As institutional births provide an opportunity for newborns to receive the initial doses of vaccines [27]. Moreover, these settings raise maternal awareness of the importance of immunization, appropriate timing for vaccines, and potential side effects, which can enhance adherence to immunization schedules [23]. Similarly, possession of a vaccination card proved crucial for vaccination coverage. Across all analyzed countries, children whose mothers reported having a vaccination card had higher coverage compared to those whose mothers did not [2, 28, 29]. This is likely because vaccination cards help caregivers track immunization schedules, encouraging full adherence.

Another important factor was attendance at prenatal care visits. Children whose mothers participated in prenatal care had higher vaccination coverage compared to those whose mothers did not receive such care. These findings align with previous research [18, 25], likely due to the guidance provided to mothers during prenatal visits regarding the importance of childhood immunization and adherence to recommended schedules [30].

This study has some limitations. The cross-sectional nature of the data does not allow for causal relationships to be established between full vaccination coverage and explanatory variables. Additionally, vaccination status relied on vaccination cards or maternal recall, which may introduce recall bias, and supply-side factors influencing immunization could not be evaluated due to data limitations. Nevertheless, the study utilized extensive, nationally representative data applicable to children aged 12–23 months in the ten analyzed countries. It also included a broad range of variables in the multivariate analysis, reinforcing the validity of the findings. Caution is recommended when generalizing results to other regions, as this study focused on ten Sub-Saharan African countries.

## Conclusion

The study identified significant variations in full vaccination coverage rates among the analyzed countries, with most exhibiting concerningly low rates. Factors such as residence, maternal education and age, economic status, place of birth, possession of a vaccination card, and frequency of prenatal care visits were identified as key determinants of vaccination coverage. These findings highlight the urgent need for integrated policies promoting maternal education, improving economic conditions, and expanding healthcare infrastructure, particularly in vulnerable areas. Additionally, targeted and accessible interventions are essential to ensure that all children have access to essential immunization, promoting health and reducing inequalities.

## Declarations

### Ethical Consideration and consent to participate

As this study was based on a secondary analysis of data from surveys publicly available through the DHS program, it was not necessary to obtain ethical approval or consent from participants for this particular study. Permission was requested from the DHS program, and authorization was granted for downloading and using the data available at http://www.dhsprogram.com. The data files do not contain names of individuals or residential addresses, ensuring the confidentiality and anonymity of the information.

### Informed Consent Statement

Not applicable

## Acknowledgments

The authors would like to acknowledge www.dhsprogram.com (access date: 18th May 2024) for giving us access to all datasets.

## Consent for publication

Not applicable.

## Availability of data and materials

In this study, publicly available datasets (DHS and MIS, between 2017 and 2022) were analyzed. Upon registration and request, these data can be found at the following link: https://dhsprogram.com/data/dataset_admin/index.cfm.

## Competing interests

The authors declare that there are no competing interests.

## Funding

This research did not receive any specific grant from funding agencies in the public, commercial, or nonprofit sectors.

## Authors’ contributions

All authors contributed to the study’s conceptualization and reviewed and submitted the manuscript. SX conducted data analysis, while SX, AV, MM, PHC, and AS were involved in interpreting and appropriating the manuscript. SX was responsible for data requests. All authors read and approved the final manuscript.

## Notes

### Competing Interest Statement

The authors have declared no competing interest.

### Author Declarations

This study utilized publicly available secondary datasets from the Demographic and Health Surveys (DHS) Program and Malaria Indicator Surveys (MIS) conducted between 2017 and 2022. Ethical approval and informed consent for data collection were obtained by the DHS Program from the relevant national institutions and participants prior to the original data collection. For this analysis, no additional ethical approval was required as the data are anonymized and publicly accessible upon registration and request through the DHS Program's platform: https://dhsprogram.com/data/dataset_admin/index.cfm.

